# Magnetocardiography can detect ventricular arrhythmia after myocardial ischemia

**DOI:** 10.1101/2025.11.04.25339469

**Authors:** Negin Ghahremani Arekhloo, Hossein Parvizi, Maryam Mardani, Agata K. Sularz, Mohamad Alkhouli, Hadi Heidari, Kianoush Nazarpour

## Abstract

**Background:** Ventricular tachyarrhythmias (VTAs) are a leading cause of sudden cardiac death after myocardial infarction (MI). Accurate risk stratification remains a clinical challenge, prompting interest in magnetocardiography (MCG) as a non-invasive diagnostic tool.

**Objective:** This systematic review aimed to evaluate the diagnostic utility of MCG in predicting post-MI VTAs, with particular attention to waveform morphology and variability.

**Methods:** We searched the PubMed, Scopus, Web of Science, and other databases for studies investigating the role of MCG in post-MI arrhythmic risk assessment, published until March 2025. Eighteen eligible studies were identified and synthesised.

**Results:** MCG-derived parameters, including temporal and spatial repolarisation heterogeneity (seven studies), late field characteristics (seven studies), and intra-QRS fragmentation (seven), were consistently reported as potential predictors of VTAs. However, most studies were limited by small cohorts, heterogeneous methodologies, and the absence of standardised diagnostic criteria.

**Conclusion:** MCG shows promise as a predictive modality for post-MI VTAs, though its clinical utility is currently constrained by methodological and scalability issues. Future developments in portable, low-cost sensors, large-scale datasets, AI-driven analytics, and quantum technologies may support wider clinical adoption and enable community-based monitoring, potentially transforming MCG into a practical tool for personalised, preventive cardiovascular care.

## 1 Introduction

Ventricular tachyarrhythmias (VTA) represent a critical and life-threatening complication frequently observed in patients after myocardial infarction [1]. The occurrence of VTA poses a significant medical challenge due to its association with a substantial proportion of sudden cardiac death [1]. Timely interventions such as revascularisation and drug therapy following myocardial ischemia (MI), including anti-platelets, statins, angiotensin converting enzyme (ACE)-inhibitors and beta-blockers, have markedly reduced the incidence of VTA [2–6]. However, despite these interventions, approximately 10% of survivors remain at high risk of sudden cardiac death in the months or years following hospital discharge, with a two-year mortality rate exceeding 25% [7]. Beyond this period, post-MI sudden cardiac death continues to pose a significant global health challenge. The development of new therapeutic strategies, such as implantable cardioverter-defibrillators (ICD), underscores the importance of effective risk stratification to identify patients with a higher likelihood of VTA following MI [8].

Although several methods for post-MI risk assessment have been proposed, their limited positive predictive value or inability to effectively guide therapy has restricted their clinical adoption [9]. For example, signal-averaged electrocardiography (SAECG) offers a high negative predictive value (NPV), but suffers from low positive predictive value (PPV) and is unsuitable for patients with bundle branch blocks [10]. Similarly, while T-wave alternans show potential for risk stratification, their predictive performance diminishes in post-MI patients on beta-blockers and is not applicable in cases of atrial fibrillation [11].

In recent years, magnetocardiography (MCG) has gained increasing recognition as a promising diagnostic method for cardiovascular diseases. MCG is a non-invasive technique that measures the cardiac magnetic field generated by the electrical currents in the heart offering several advantages over traditional electrocardiography (ECG). Unlike ECG, MCG is less influenced by conductivity of the surrounding tissues, such as lungs, bones, fat, and skin, which are largely transparent to magnetic fields. MCG also demonstrates greater sensitivity to tangential currents, often associated with myocardial scar tissue, and to circular currents that remain undetected by ECG, thereby enhancing its ability to detect subtle electrophysiological abnormalities. Furthermore, as MCG requires no skin contact, it avoids issues related to contact resistance and muscle motion artifacts, enabling faster, simultaneous acquisition of signals from multiple locations.

To date, numerous studies have supported the use of MCG for identifying patients at high risk of post-MI VTAs [9, 12, 13]. These investigations have primarily utilised superconducting quantum interference devices (SQUIDs), typically operated within magnetically shielded rooms to suppress environmental electromagnetic noise from nearby electronic equipment. However, recent technological advancements now enable reliable magnetic recordings in unshielded environments by employing second- or higher-order gradiometer configurations and/or active real-time electronic noise cancellation techniques [14]. These innovations significantly reduce the need for heavy electromagnetic shielding, paving the way for broader clinical adoption of MCG, including in routine hospital settings and interventional procedures [14, 15]. In parallel, several types of novel, non-cryogenic magnetic sensors, such as optically pumped magnetometers, magnetoresistive sensors, and induction coils, have emerged [16, 17]. These devices are portable, operate at room temperature, and can function with minimal or no electromagnetic shielding, making bedside application feasible. Furthermore, if the cost of these sensors continues to decrease and integration with user-friendly interfaces and remote monitoring systems improves, community-based care and wider outpatient deployment of MCG could become a practical and scalable reality.

Despite advancements, our research reveals a notable gap: the absence of comprehensive systematic reviews or meta-analyses evaluating MCG’s effectiveness in detecting VTA risk in post-MI patients. We assert that a rigorous and systematic assessment of this diagnostic approach, especially in the contexts of waveform morphology and signal variability analysis, is urgently needed to guide clinicians and support evidence-based adoption in cardiovascular care.

Accordingly, this review was undertaken to evaluate the diagnostic utility of MCG for VTA risk stratification in post-MI patients, with a particular focus on waveform parameters and signal variability. In this context, four categories of MCG features were considered: temporal dispersion of ventricular repolarisation inhomogeneity, spatial dispersion of ventricular repolarisation inhomogeneity, time-domain late-field analysis, and intra-QRS fragmentation. We show that MCG waveform parameters, such as repolarisation heterogeneity, late field characteristics, and intra-QRS fragmentation, hold significant potential for predicting post-MI VTAs.

## 2 Methodology

This systematic review was conducted in accordance with established guidelines for evidence synthesis (PRISMA). A comprehensive search was performed across PubMed (including MEDLINE complete), academic search ultimate, CINHAL Ultimate, AMED, Embase, the Cochrane Central Register of Controlled Trials, and ClinicalTrials.gov, covering studies published up to March 2025.

The search strategy combined terms relating to the intervention, outcomes, and population of interest. Intervention-related terms included “magnetocardiography”, “magnetocardiogram”, and “MCG”. Outcomes were described using the terms “ventricular tachycardia”, “VT”, “ventricular tachyarrhythmia”, “VTA”, “ventricular arrhythmia”, and “VA”. Population terms included “ischaemic heart disease”, “ischemic heart disease”, “myocardial infarction”, “myocardial ischemia”, “myocardial ischaemia”, “IHD”, “MI”, “coronary artery disease”, “CAD”, “coronary disease”, “cardiovascular disease”, “CVD”, “coronary heart disease”, “CHD”, “stroke”, and “heart stroke”.

### 2.1 Study selection

Two independent reviewers (NGA and HP) conducted an initial screening by evaluating the titles and abstracts of retrieved studies. This was followed by a detailed assessment of the full-text articles for studies deemed potentially eligible, to confirm their inclusion in the systematic review. Any discrepancies between reviewers regarding study inclusion were resolved through discussion and consensus, with input from senior authors sought when agreement could not be reached. Reference lists of the included studies were also screened to identify any potentially relevant articles that might have been missing during the initial screening.

Studies were included if they were cross-sectional or case–control in design, published in English, and involved adult participants (≥18 years) with ventricular tachycardia and a history of ischaemic or coronary heart disease, specifically focusing on those reporting diagnostic outcomes of MCG based on morphological changes in the MCG waveform. Animal studies, case reports, reviews, conference abstracts, letters to the editor, and editorials were excluded. Research only consists of ‘completed studies’ and those ‘with results’.

### 2.2 Data Extraction

From each included study, the following attributes were extracted: publication details, cohort size and characterisation (underlying etiology), study design (retrospective/prospective), type of MCG technology, number of channels, MCG features studied, use of magnetic field shielding, and study limitation, as listed in Table 1. The methodological quality of included studies was evaluated independently by two reviewers using the Quality Assessment of Diagnostic Accuracy Studies 2 (QUADAS-2) tool. Details of the risk-of-bias and applicability concern assessment are provided in Table 2 and Table 3, respectively.

**Table 1:** Summary of included trial design, significant findings and limitations of each trial. All trials used SQUID inside the shielded environment.

| Author & Year | Total number of patients | Patients with VA | Patients with CVD and no VA | Healthy controls | Retrospective /Prospective | Number of MCG channels [SQUID] | Significant findings | Limitations |
| --- | --- | --- | --- | --- | --- | --- | --- | --- |
| Hailer et al. (1998) [18] | 28 | 8 | 10 | 10 | Retrospective | 36 | 1. QTd (QRS, JTa, TaT, QTa, QT) is greater in the patient group compared to healthy, but the difference between pts w/wo MI was not significant.<br>2. SI and SIn can differentiate pts w/wo VT. SI/ SIn for **JTa and QTa** were significantly higher in Pts with VT. | 1. Small sample size.<br>2. some patients were on Metoprolol (B-Blocker).<br>3. Lower EF in the VT patients.<br>4. Retrospective |
| Oikarinen (1998) [19] | 18 | 10 | 8 | - | Retrospective | Single channel from 42 locations | 1. Mean QRS duration is much longer in VT patients compared to MI without VT. | 1. 5 patients in the VT group were on antiarrhythmic.<br>2. EF is lower in VT patients.<br>3. The follow-up time after MI in patient group is different.<br>4. EPS study had been done just in VT group.<br>5. Retrospective |
| Oikarinen (2001) [20] | 73 | 32 | 28 | 13 | Retrospective | 33 | 1. QRS duration and T wave peak to T wave end (TPE) are greater in VT patients compared to MI patients without VT.<br>2. Mean TPE and mean filtered QRS are **independently** associated with VT susceptibility. | 1. Different number of patients in two group were on betablocker.<br>2. More LV aneurysm in the VT patients.<br>3. Retrospective |
| Mäkijärvi 1993 [13] | 30 | 10 | 10 | 10 | Retrospective | Single channel MCG from 9 locations | 1. fQRS is prolonged in VT group compared to the MI group.<br>2. Smaller RMS value (especially last 60 ms) compared to the MI group.<br>3. prolonged low-amplitude signal duration in VT group compared to the MI group | 1 lower EF from VT patient group ($p>0.05$).<br>2. Higher number of MI attacks in Vt patients ($p<0.05$)<br>3. Retrospective |
| Korhonen 2000 [9] | 100 | 38 | 62 | - | Retrospective | 7 | QRS duration, RMS of late fields (last 60,40 ms: RMS<sub>40</sub> and RMS<sub>60</sub>), and low-amplitude signal duration below 300 and 500 fT (LAS<sub>300</sub> and LAS<sub>500</sub>) were different in VT patients. | 1. Short follow-up time (6 months) after MI.<br>2. Higher rate of aneurysm in the VT group (p<0.05).<br>3. Retrospective |
| Korhonen 2001[21] | 136 | 53 | 83 |  | Retrospective | 7 | Fragmentation score (s), QRS duration, RMS of late fields (RMS<sub>40</sub> and RMS<sub>30</sub>), and LAS<sub>300</sub> and LAS<sub>500</sub> discriminate VT patients from non-VT patients. | 1. Short follow-up time (6 months) after MI.<br>2. Higher rate of aneurysm in the VT group (p<0.05).<br>3. Retrospective |
| Korhonen 2002 [22] | 22 | 22 | 22 | - | Prospective | 7 | QRS duration, RMS<sub>40</sub>, LAS<sub>300</sub>, Intra-QRS fragmentation (M and S) were strongly higher in VT patients before surgery. The correlation of intra-QRS fragmentation to the end of ventricular excitation were stronger, compared to time-domain parameters. | Evaluation has been done within a short duration after surgery. |
| Korhonen 2006 [23] | 158 | 18 | 140 |  | Prospective | 7 | Fragmentation score is higher in the VT patients (p=0.006) | 1. Patient population is small.<br>2. short period after MI, so not including remote MI. |
| Korhonen 2002 [24] | 44 | 22 | 22 | - | Retrospective | 7 | Significant difference in QRSD, RMS<sub>40</sub>, and LAS<sub>300</sub> has been observed between two patient groups. | 1. More LV aneurysm in VT patients (p<0.05)<br>2. small number of participants<br>3. Retrospective |
| Korhonen 2006 [25] | 158 | 18 | 140 |  | Prospective | 7 | longer QRS duration predicts the VT propensity after MI. | 1. No continuous monitoring for the VT patients.<br>2. Relatively small number of the patients |
| Endt, P. 2000 [26] | 31 | 20 | 11 | - | Retrospective | Single channel MCG from 9 locations | Intra-QRS fragmentation scores (M and S scores) in the whole QRS duration is useful for differentiating VT pts from MI. | 1. Significant lower EF in the VT group (p<0.05).<br>2. Relatively small number of patients. |
|  |  |  |  |  |  |  |  | 3. ECG and MCG measurements were made sequentially rather than simultaneously.<br>4. Retrospective |
| Göudde 2001 [27] | 119 | 43 | 42 | 34 | Retrospective | 49 | Intra-QRS fragmentation score can identify patients at risk of malignant arrhythmia. | 1. Retrospective VT patient selection can cause bias.<br>2. small number of patients.<br>3. Retrospective |
| Mäkijärvi 1993 [28] | 33 | 11 | 11 | 11 | Retrospective | Single channel MCG from 9 locations | fQRS and RMS duration can separate VT patient from MI patients. | 1. ECG and MCG recordings have not been done simultaneously.<br>2. Small group<br>3. Retrospective |
| Weismüller 1993 [29] | 12 | 4 | 4 | 4 | Retrospective | 37 | QRS duration, RMS, and LAS are not able to discriminate patients with VT susceptibility. | 1. very small size<br>2. Retrospective |
| Müller 1999 [30] | 119 | 43 | 42 | 34 | Retrospective | 49 | QRS duration and Fragmentation score can separate patients with and without propensity to VT. | 1. Short follow-up time after MI.<br>2. Retrospective<br>3. Detail of the patient group are not mentioned. |
| Leeuwen 2003 [31] | 85 | 11 | 54 | 20 | Retrospective | 36 | QTd and SI can discriminate VT from MI patients. | 1. Retrospective<br>2. Small number |
| Van Leeuwen 2006 [32] | 144 | 15 | 79 | 50 | Retrospective | 37 | QTd and SI can discriminate VT from MI patients. | 1. Retrospective<br>2. Small number<br>3. Detail of the patient group are not mentioned |
| Schless 2003 [33] | 50 | 10 | 25 | 15 | Retrospective | 55 | QRS microvariability is not able to discriminate between MI and VT patients | 1. small number of patients |

**Table 2:**
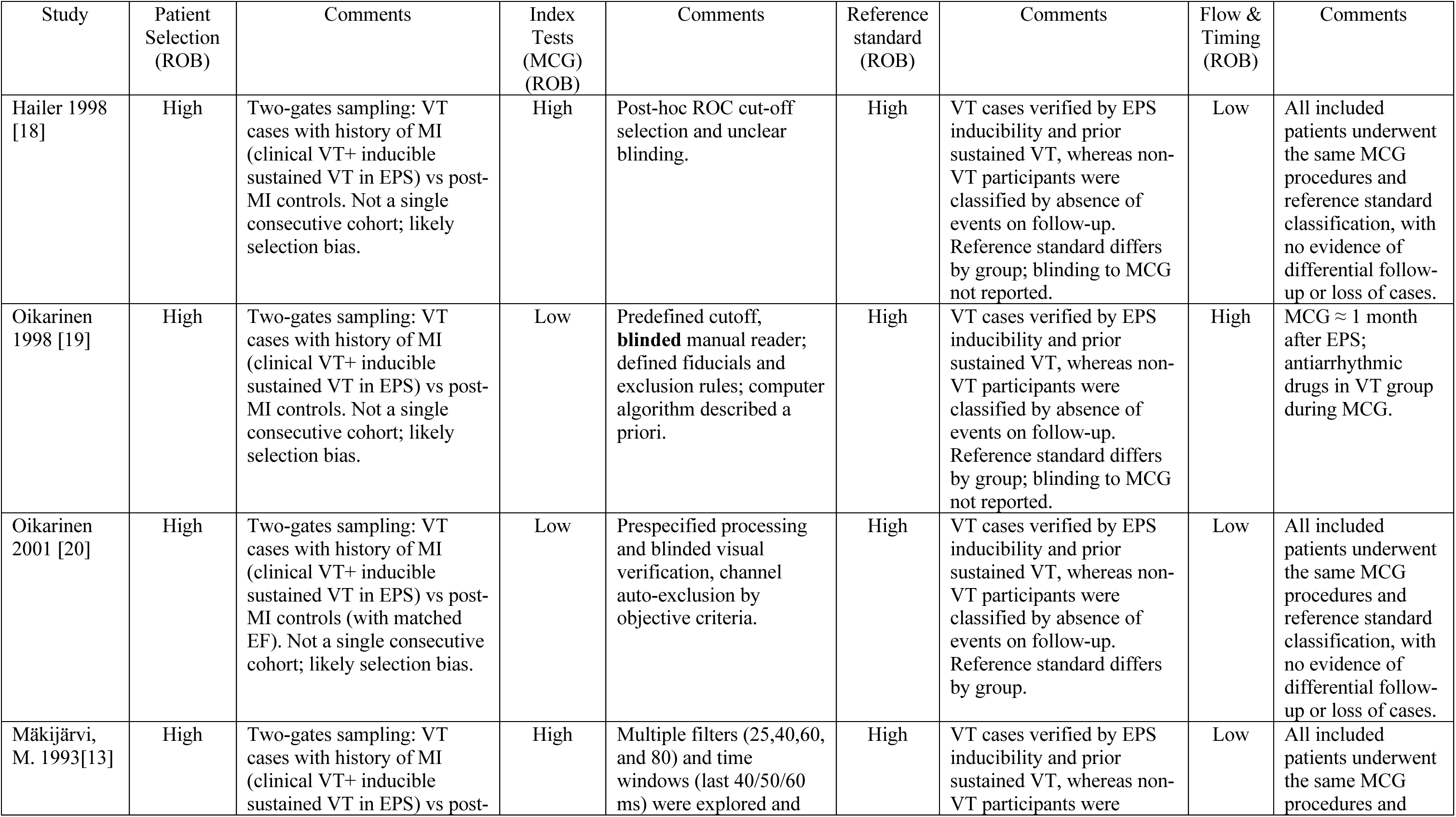
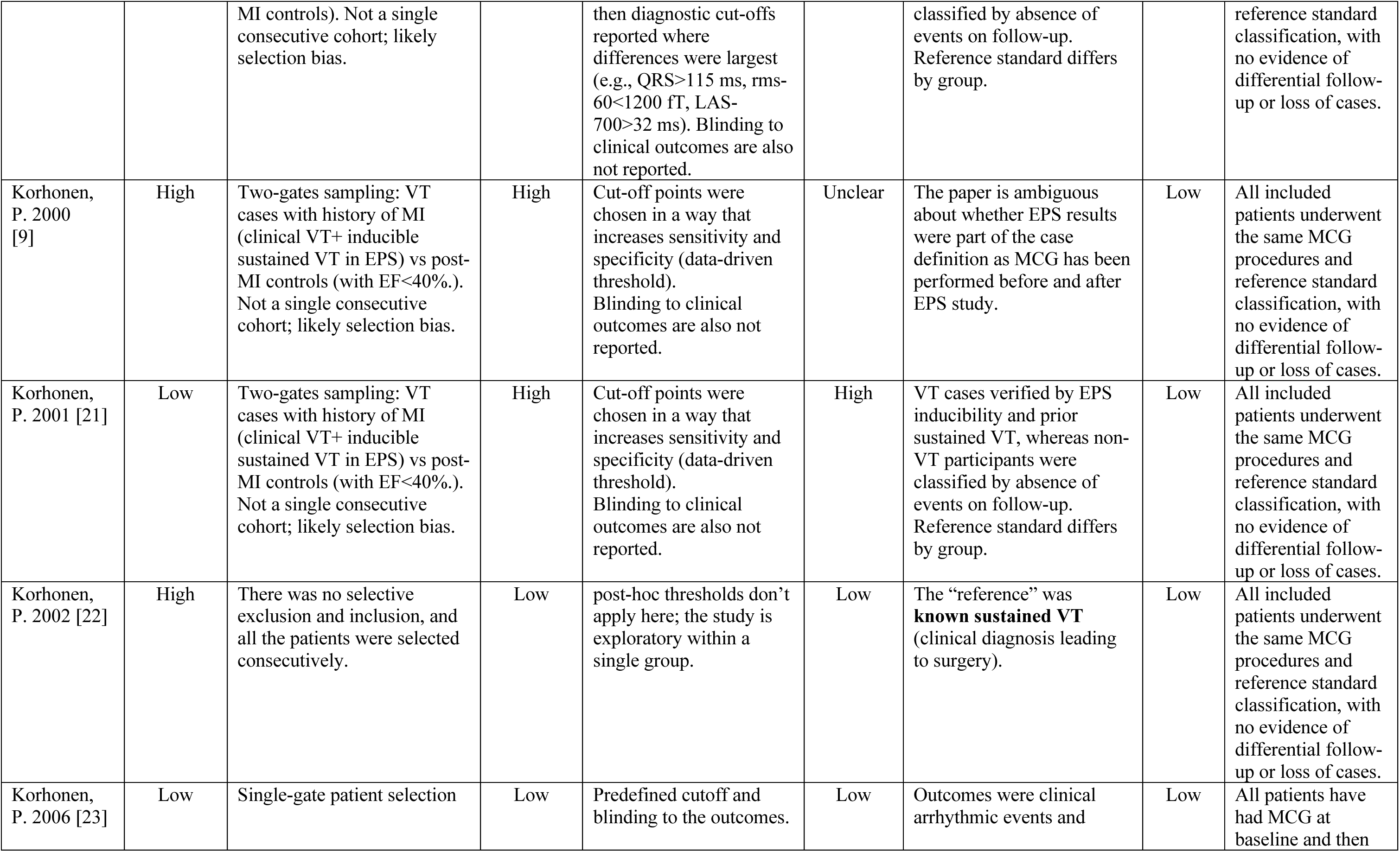
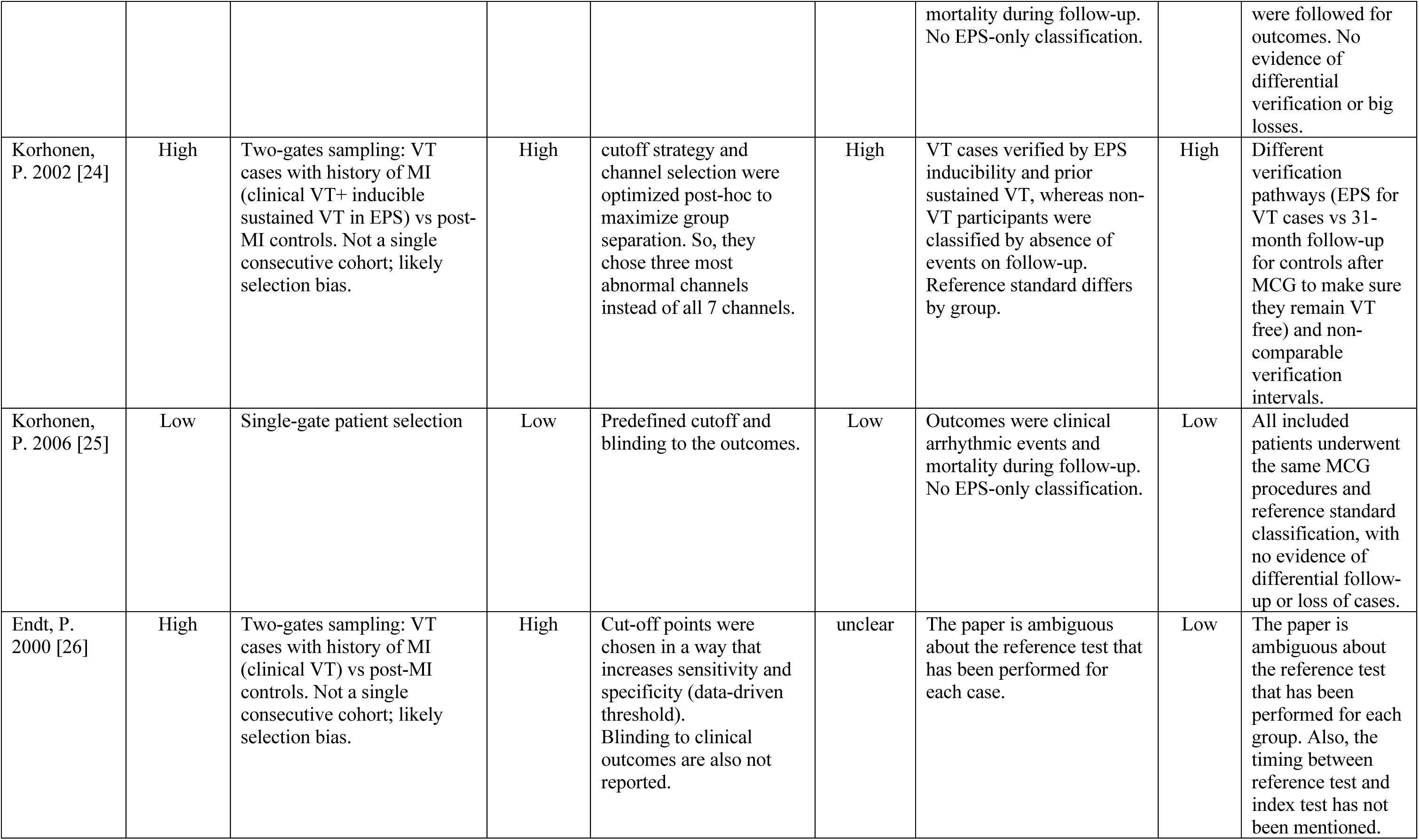
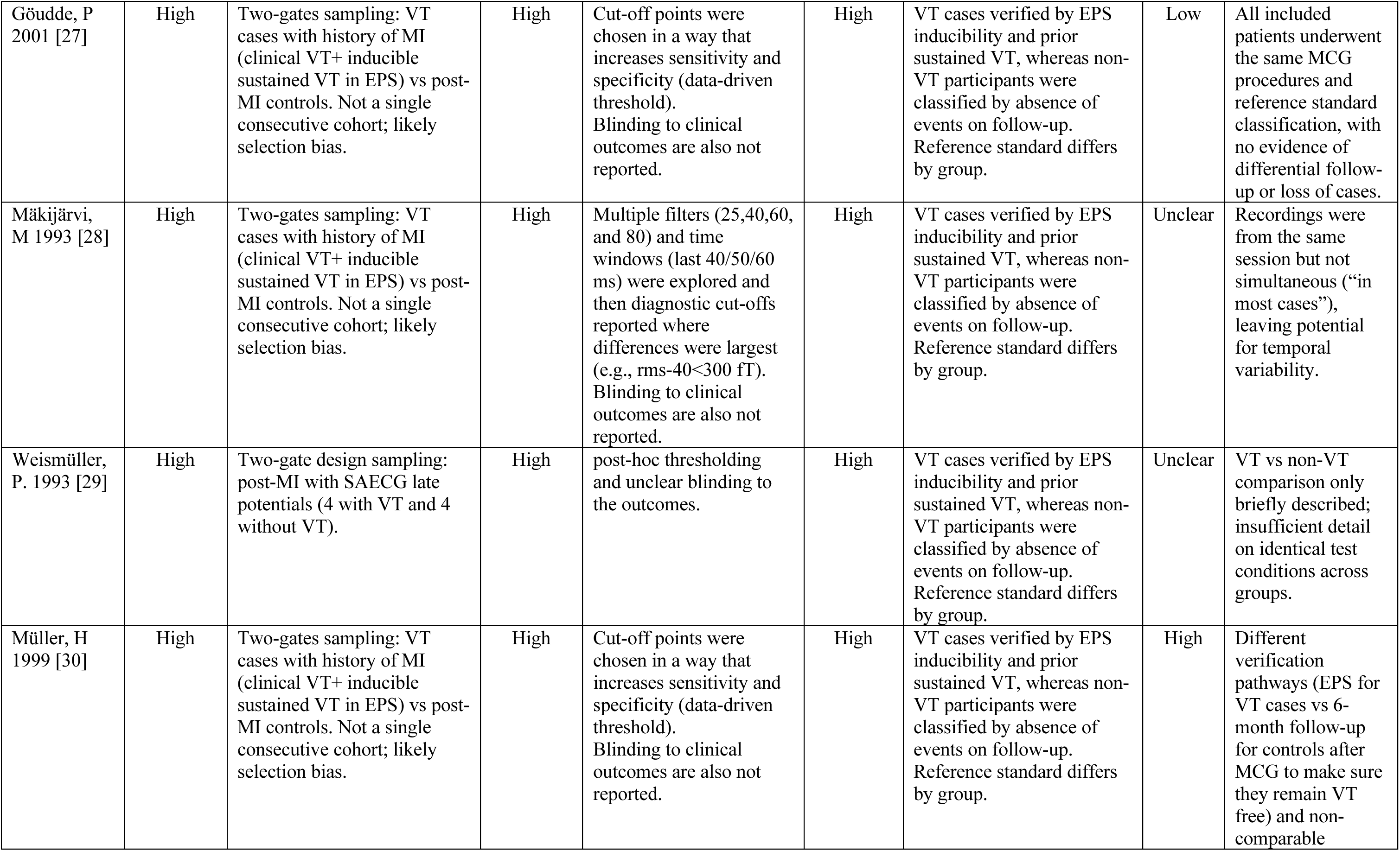
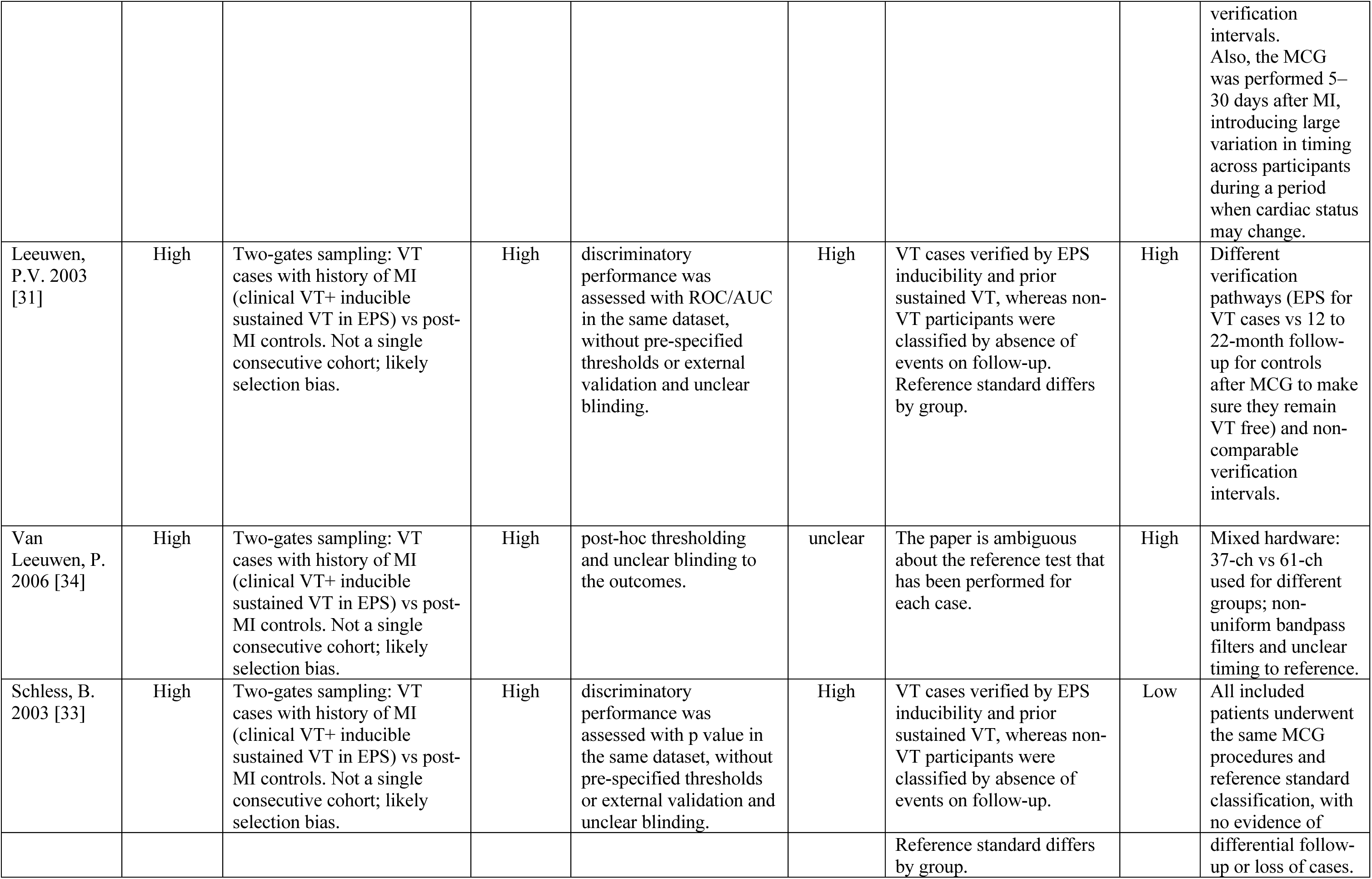
Risk of Bias assessment using Quality Assessment of Diagnostic Accuracy Studies 2 tool (QUADAS-2)

**Table 3:** Applicability Concern using Quality Assessment of Diagnostic Accuracy Studies 2 tool (QUADAS-2)

| Study | Patient Selection | Comments | Index Tests (MCG) | Comments | Reference standard | Comments |
| --- | --- | --- | --- | --- | --- | --- |
| Hailer 1998 [18] | Low | Although the use of different verification pathways (EPS/clinical VT vs. event-free follow-up) introduces potential risk of bias in patient selection, this approach reflects clinical practice, where not all post-MI patients undergo EPS. | Low | Index tests (Dispersion and smoothness measures) align with the parameters in our question and, they try to answer the same disorder as us. | Low | Despite verification differences, these definitions are broadly aligned with clinical practice for post-MI arrhythmic risk. |
| Oikarinen 1998 [19] | Low | Although the use of different verification pathways (EPS/clinical VT vs. event-free follow-up) introduces potential risk of bias in patient selection, this approach reflects clinical practice, where not all post-MI patients undergo EPS. | Low | Index tests (QT interval dispersion) align with the parameters in our question and, they try to answer the same disorder as us. | Low | Despite verification differences, these definitions are broadly aligned with clinical practice for post-MI arrhythmic risk. |
| Oikarinen 2001 [20] | High | Controls were matched to VT cases by left ventricular EF, which does not reflect our target population across the full EF spectrum; this limits applicability of the findings | Low | Index tests (QT interval and QRS duration) align with the parameters in our question and, they try to answer the same disorder as us. | Low | Despite verification differences, these definitions are broadly aligned with clinical practice for post-MI arrhythmic risk. |
| Mäkijärvi, M. 1993[13] | Low | Although the use of different verification pathways (EPS/clinical VT vs. event-free follow-up) introduces potential risk of bias in patient selection, this approach reflects clinical practice, where not all post-MI patients undergo EPS. | Low | Index tests (QRS duration, RMS, and LAS) align with the parameters in our question and, they try to answer the same disorder as us. | Low | Despite verification differences, these definitions are broadly aligned with clinical practice for post-MI arrhythmic risk. |
| Korhonen, P. 2000 [9] | High | Controls were limited to LVEF $\leq 0.40$ to match VT cases, which does not reflect our target population across the full EF spectrum; enrolment changed from consecutive to selective. | Low | Index tests (QRS duration, RMS, and LAS) align with the parameters in our question and, they try to answer the same disorder as us. | Low | Despite verification differences, these definitions are broadly aligned with clinical practice for post-MI arrhythmic risk. |
| Korhonen, P. 2001 [21] | High | Controls were limited to LVEF $\leq 0.40$ to match VT cases, which does not reflect our target population across the full EF spectrum; enrolment changed from consecutive to selective. | Low | Index tests (fragmentation, QRS duration, RMS, and LAS) align with the parameters in our question and also, they try to answer the same disorder as us. | Low | Despite verification differences, these definitions are broadly aligned with clinical practice for post-MI arrhythmic risk. |
| Korhonen, P. 2002 [22] | High | Surgical cohort of known sustained VT/VF post-MI (not a risk-stratification population); not representative. | Low | Index tests (fragmentation, QRS duration, RMS, and LAS) align with the parameters in our question and also, they try to answer the same disorder as us. | Low | The “reference” was **known sustained VT** (clinical diagnosis leading to surgery). |
| Korhonen, P. 2006 [23] | Low | Although the use of different verification pathways (EPS/clinical VT vs. event-free follow-up) introduces potential risk of bias in patient selection, this approach reflects clinical practice, where not all post-MI patients undergo EPS. | Low | Index tests (fragmentation, QRS duration) align with the parameters in our question and, they try to answer the same disorder as us. | Low | Despite verification differences, these definitions are broadly aligned with clinical practice for post-MI arrhythmic risk |
| Korhonen, P. 2002 [24] | Low | Although the use of different verification pathways (EPS/clinical VT vs. event-free follow-up) introduces potential risk of bias in patient selection, this approach reflects clinical practice, where not all post-MI patients undergo EPS. | Some concerns | Using “3 most abnormal channels” introduces a subjective/retrospective element that doesn’t easily generalize. | Low | Despite verification differences, these definitions are broadly aligned with clinical practice for post-MI arrhythmic risk. |
| Korhonen, P. 2006 [25] | Low | These patients resemble the population of our systematic review (post-MI, risk stratification) | Low | Index test (QRS duration) align with the parameters in our question and, they try to answer the same disorder as us. | Low | Clinical outcomes directly match the review’s focus. |
| Endt, P. 2000 [26] | Low | Although the use of different verification pathways (EPS/clinical VT vs. event-free follow-up) introduces potential risk of bias in patient selection, this approach reflects clinical practice, where not all post-MI patients undergo EPS. | Low | Index test (Fragmentation score, QRS duration, LAS, and RMS) align with the parameters in our question and, they try to answer the same disorder as us. | Low | Despite verification differences, these definitions are broadly aligned with clinical practice for post-MI arrhythmic risk. |
| Göudde, P 2001 [27] | Low | Although the use of different verification pathways (EPS/clinical VT vs. event-free follow-up) introduces potential risk of bias in patient selection, this approach reflects clinical practice, where not all post-MI patients undergo EPS. | Low | Index test (Fragmentation score) align with the parameters in our question and, they try to answer the same disorder as us. | Low | Despite verification differences, these definitions are broadly aligned with clinical practice for post-MI arrhythmic risk. |
| Mäkijärvi, M 1993 [28] | Low | Although the use of different verification pathways (EPS/clinical VT vs. event-free follow-up) introduces | Low | Index test (QRS duration and RMS) align with the parameters in our question | Low | Despite verification differences, these definitions are broadly aligned with clinical practice for post-MI arrhythmic risk. |
|  |  | potential risk of bias in patient selection, this approach reflects clinical practice, where not all post-MI patients undergo EPS. |  | and, they try to answer the same disorder as us. |  |  |
| Weismüller, P. 1993 [29] | High | selection was based on presence of SAECG late potentials as well which doesn't represent the intended population. | Low | Index test (QRS duration, LAS, and RMS) align with the parameters in our question and, they try to answer the same disorder as us. | Moderate | Outcomes (sustained VT vs. no VT) are clinically relevant, but the restriction to late-potential positive patients narrows applicability. |
| Müller, H 1999 [30] | Low | Although the use of different verification pathways (EPS/clinical VT vs. event-free follow-up) introduces potential risk of bias in patient selection, this approach reflects clinical practice, where not all post-MI patients undergo EPS. | Low | Index test (QRS duration, and fragmentation score) align with the parameters in our question and, they try to answer the same disorder as us. | low | Despite verification differences, these definitions are broadly aligned with clinical practice for post-MI arrhythmic risk. |
| Leeuwen, P.V. 2003 [31] | Low | Although the use of different verification pathways (EPS/clinical VT vs. event-free follow-up) introduces potential risk of bias in patient selection, this approach reflects clinical practice, where not all post-MI patients undergo EPS. | Low | Index test (QRS duration, SI, SIn) align with the parameters in our question and, they try to answer the same disorder as us. | low | Despite verification differences, these definitions are broadly aligned with clinical practice for post-MI arrhythmic risk. |
| Van Leeuwen, P. 2006 [34] | Low | Although the use of different verification pathways (EPS/clinical VT vs. event-free follow-up) introduces potential risk of bias in patient selection, this approach reflects clinical practice, where not all post-MI patients undergo EPS. | Low | Index test (QT dispersion quantified with SI, SIn) align with the parameters in our question and, they try to answer the same disorder as us. | High | Mixed hardware: 37-ch vs 61-ch used for different groups; non-uniform bandpass filters. |
| Schless, B. 2003<br>[33] | Low | Although the use of different verification pathways (EPS/clinical VT vs. event-free follow-up) introduces potential risk of bias in patient selection, this approach reflects clinical practice, where not all post-MI patients undergo EPS. | Low | Index test (QRS microvariability) align with the parameters in our question and, they try to answer the same disorder as us. | low | Despite verification differences, these definitions are broadly aligned with clinical practice for post-MI arrhythmic risk. |

### 2.3 Study flow

As illustrated in Figure 1, a total number of 276 studies were initially identified through comprehensive database searches.

**Figure 1:**
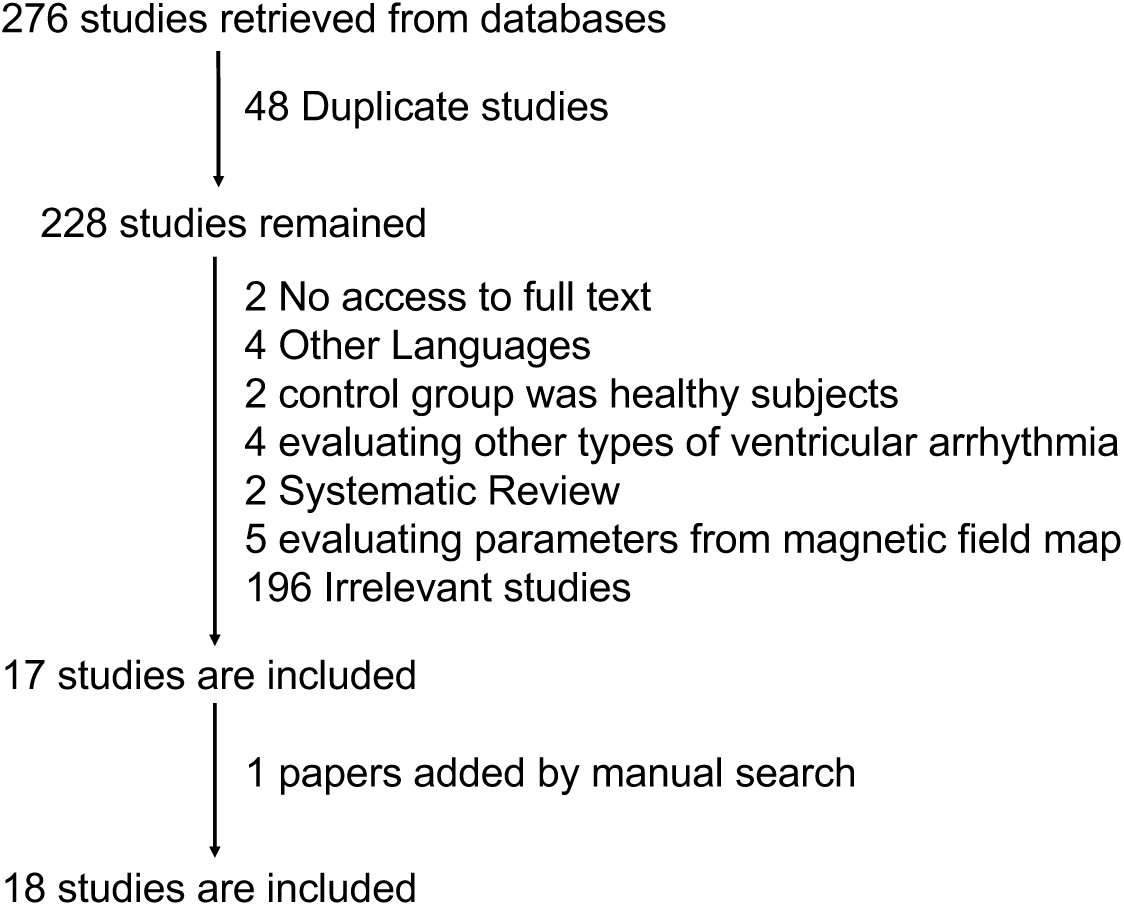
Prisma Chart.

**Figure 2:**
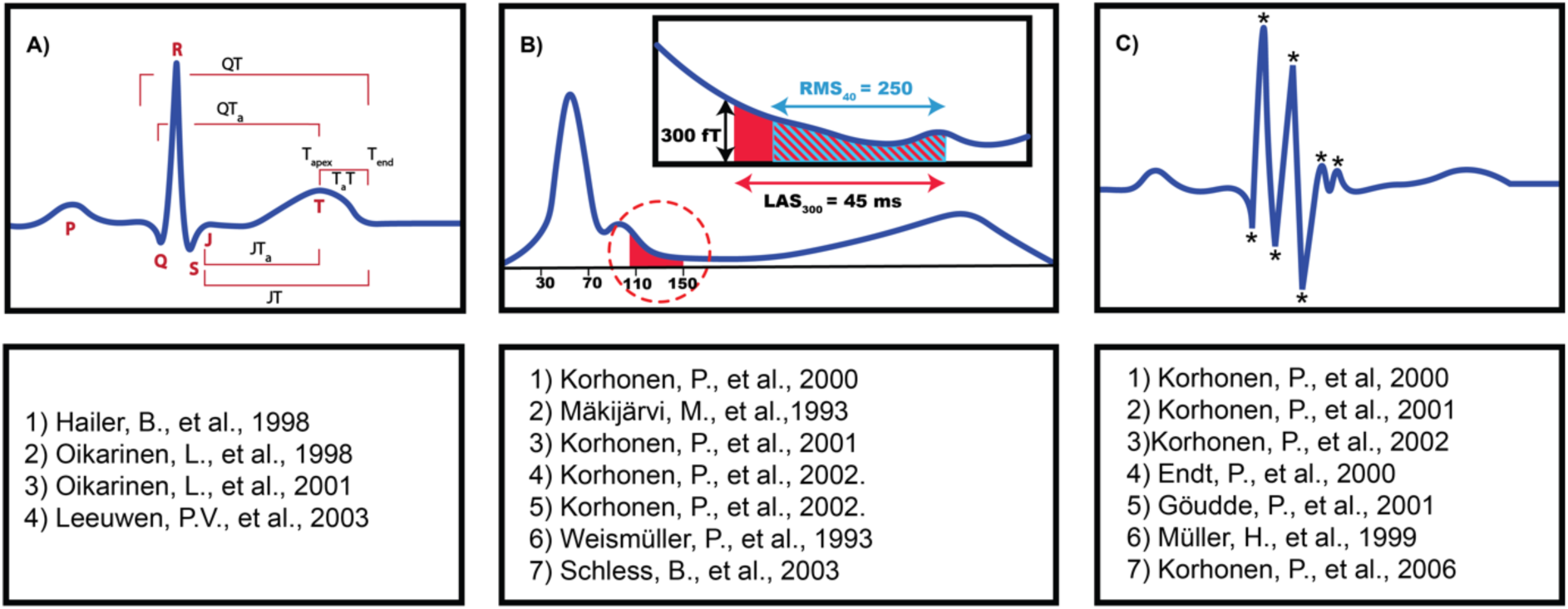
MCG waveform attributes: A) Repolarisation parameters B) Time-domain late field analysis C) Intra-QRS fragmentation.

After removal of 48 duplicates, 228 studies remained for the screening. Of these, 196 papers were excluded at the abstract stage for not focusing on arrhythmic risk. At full-text review, a further 19 articles were excluded due to: lack of access to the full text (n=2), non-English language (n=4), use of healthy control groups only (n=2), systematic reviews (n=2), evaluation of magnetic field map parameters rather than waveform morphology (n=5), and assessment of non-target ventricular arrhythmias (n=4). Therefore, Seventeen studies were included, and an additional one study was identified through manual reference searching, resulting in a total of 18 studies included in the final review.

### 2.4 Outcome of interest

Four categories of MCG waveform parameters were prespecified for extraction and synthesis: (1) temporal dispersion of ventricular repolarisation inhomogeneity, (2) spatial dispersion of ventricular repolarisation inhomogeneity, (3) time-domain late-field analysis, and (4) intra-QRS fragmentation analysis (see **Error! Reference source not found.**).

The included studies in this review were mainly exploratory, with non-standardised methodologies of MCG processing and frequent reliance on post-hoc thresholding. Also, the majority of studies (15/18) identified VTAs retrospectively, enrolling patients only after an arrhythmic event. This approach excludes individuals who did not survive and reflects retrospective patient selection. Prospective studies were less common, limiting the consistent capture of new arrhythmic events during follow-up. Taken together, substantial variation in methodology, data reporting, and analytic strategies necessitates a narrative reporting with tabulation of key study features.

## 3 Result

### 3.1 Temporal Dispersion of Ventricular Repolarisation Inhomogeneity

Myocardial ischemia leads to formation of scar tissue, which disrupts the heart’s normal electrical pathways [35]. This disruption slows electrical conduction and increases repolarisation heterogeneity across the damaged myocardium, making it more susceptible to re-entry of ventricular electrical currents and the subsequent development of ventricular tachyarrhythmia [36]. Irregularities in repolarisation timing are reflected in various QT-related intervals, including:

● QTend (from Q onset to T wave end: QT),
● QTapex (from Q onset to T wave apex: QTa),
● JTapex (from J onset to T wave apex: JTa),
● JTend (from J onset to T wave end: JT), and
● TaT (from T apex to T wave end: TapexTend).

Recent studies suggest that the risk of ventricular arrhythmias is more closely associated with the temporal dispersion of these intervals rather than their absolute durations. Temporal dispersion refers to the difference between the maximum and minimum values recorded across leads. The greater the temporal dispersion, the higher the vulnerability to VTA occurrence. Among these intervals, increased QT dispersion (QTd) has been consistently linked to the VTAs in multiple studies, establishing it as a significant predictor of VTA risk [37, 38].

Within the scope of this systematic review, four out of eighteen studies investigated QTd as a criterion to distinguish between MI patients susceptible to VTAs and those who are not [18–20, 31]. Out of these, two studies reported a significant association between increased QTd and higher susceptibility to VTAs post-MI [19, 31], while the other two did not confirm this relationship. This discrepancy may stem from the method of calculating QTd, which relies on comparing only the most extreme values, making it highly susceptible to outliers, particularly due to challenges in accurately determining the T wave offset [39].

Notably, among the studies that did not identify QTd as a predictive marker for arrhythmia risk, one reported that TaT dispersion - another parameter indicative of repolarisation inhomogeneity-could effectively distinguish patients with a predisposition to arrhythmias post-MI [20].

These results suggest that the temporal aspects of repolarisation inhomogeneity may serve as a viable predictive marker for identifying patients at risk of VTAs following MI. However, a more comprehensive understanding requires evaluating this inhomogeneity from an additional perspective. Specifically, examining repolarisation differences across adjacent MCG channels on the thorax enables a more detailed analysis of the spatial dimensions of QT variations. This approach, referred to as the spatial aspects of repolarisation heterogeneity, will be described in the following sections.

### 3.2 Spatial Dispersion of Ventricular Repolarisation Inhomogeneity

Since QTd captures only the temporal aspect and overlooks the spatial heterogeneity of repolarisation, it may not fully represent the complex nature of ventricular repolarisation. Consequently, incorporating the spatial distribution of repolarisation heterogeneity has been proposed to enhance the diagnostic accuracy in identifying post-MI patients at a higher risk of VTAs.

Spatial dispersion is assessed visually by plotting the deviation of each ECG or MCG channel from the minimum QT value relative to its spatial position. Local and global irregularities are then quantified using two smoothness indices, the smoothness index (SI) and its normalized form (SIn).

The former is calculated by first estimating the average QT variation between each channel and its adjacent channels. The SI is then derived as the mean of these variations across all channels, quantifying the spatial variation of QT values. A higher SI reflects greater repolarisation heterogeneity [40].

To account for natural variations in cardiac function, the SI is normalised, resulting in SIn. To this end, a baseline *norma*” spatial QT pattern is established using data from healthy subjects. This reference is used to assign a weighting factor to each channel, effectively adjusting for the expected inhomogeneity found in normal hearts. The SIn is then calculated using the same method as the SI but incorporating these weighting factors to correct for physiological variability [40]. This approach captures both local and global irregularities, with higher SIn values indicating greater deviation from the established normal repolarisation pattern [31].

Three out of eighteen reviewed studies specifically assessed the spatial distribution of QTd using SI and SIn, for stratify post-MI patients by VTAs risk [18, 31, 32]. All three found that SI and SIn offered superior discriminatory power compared to purely temporal measures. This advantage likely stems from their ability to capture differences in how dispersion is spatially distributed, even when the overall level of dispersion appears similar.

Multichannel MCG technology, which captures signals from multiple thoracic locations, offers a more comprehensive view of the repolarization process. Evidence indicates that broader chest coverage increases the range of observed QT durations, thereby enhancing SI and SIn values and improving their effectiveness in clinical risk stratification [31].

### 3.3 Time-domain Analysis of the Late Field

Another MCG parameter utilised in post-MI risk stratification is the time-domain analysis of the late fields, which are small deflections observed at the terminal portion of the QRS complex. These deflections reflect abnormal ventricular activation and are considered the magnetic counterparts of late potentials, which are low-amplitude, high-frequency electrical signals indicating delayed and fragmented conduction in the myocardium. Such conduction abnormalities are often associated with arrhythmogenic substrates following MI. Identifying the spatial origin of these late fields may help in localising the arrhythmic substrate, providing valuable guidance for targeted catheter ablation.

For the assessment of MCG late fields, the following indices are computed:

● QRS duration (QRSd),
● Root mean square (RMS): Amplitude of the magnetic field strength during the last 30, 40, 50, and 60 ms (known as RMS30, RMS40, RMS50, and RMS60, respectively),
● Duration of low amplitude signals (LAS) lower than 300fT, 500fT, 600fT, and 700fT (also known as LAS300, LAS500, LAS600, LAS700, respectively) (fT= 10-15 T).
● QRS duration variability

Six of the eighteen studies examined the first three indexes from the time-domain analysis of late field [9, 13, 21, 22, 24, 29]. Five of these reported that MCG late fields can effectively distinguish post-MI patients who experienced sustained VTAs from those without such arrhythmias [9, 13, 21, 22, 24]. Findings indicate that patients with post-MI VTAs show prolonged QRS and LAS durations, along with reduced RMS amplitude. While these abnormal late ventricular activities associated with VTA risk are detectable in unfiltered MCG recordings, applying filters often improves detection and enhances diagnostic accuracy.

One study was unable to distinguish between patients with and without VTAs after MI using late field indices. This discrepancy may be due to the challenge of differentiating true late QRS activity from background noise [29].

Furthermore, four studies independently evaluated QRS duration [18, 19, 25, 28]. Consistently, these studies found that patients with VTAs exhibited prolonged QRS durations compared to those without. This prolongation may serve as a valuable metric for stratifying post-MI patients into VTAs and non-VTAs subgroups.

Finally, one study [33] examined beat-to-beat variations in QRS duration and found increased QRS duration microvariability in patients with arrhythmias following MI.

### 3.4 Intra-QRS fragmentation Analysis

Intra-QRS fragmentation analysis quantifies high-frequency components within the QRS complex using binomial filtering through a non-recursive filter. The analysis is further refined by counting the number of polarity changes, or extrema, within the QRS complex. A fragmentation score is calculated to reflect the degree of intra-QRS notching, based on both the number and magnitude of extrema. The prognostic value of the fragmentation score for identifying patients at risk of VTAs post-MI was assessed in seven studies. All reported higher fragmentation scores in patients with VTAs compared to those without, highlighting its potential as a predictive marker for post-MI VTAs [9, 21–23, 26, 27, 30].

## 4 Discussion

This systematic review comprehensively evaluates the diverse MCG waveform attributes used to detect the VTAs following MI. With sudden cardiac death due to VTAs posing a significant threat to post-MI patients, there is an urgent need for more accurate and reliable risk stratification tools. MCG, as a non-invasive and contactless diagnostic method, offers significant promise by detecting subtle physiological abnormalities that often go undetected by conventional ECG.

As summarised Table 1, a substantial number of included studies in this review suggest that MCG waveform parameters, such as repolarisation heterogeneity, late field characteristics, and intra-QRS fragmentation, hold significant potential for predicting post-MI VTAs. In particular, measures such as QTd and the smoothness indices (SI/SIn) have demonstrated strong correlations with VTA risk. Additionally, time-domain analysis of the late field and intra-QRS fragmentation scores have shown promise in distinguishing high-risk post-MI patients from those with lower susceptibility to VTAs. Despite these encouraging results, several challenges remain. While the majority of studies support MCG’s clinical utility, inconsistencies persist regarding the predictive power of certain parameters, likely due to methodological variations and differences in patient populations. Hence, further validation of MCG’s diagnostic accuracy is essential, especially in larger and more diverse patient populations. Future research should also prioritise integrating MCG into routine clinical workflows and evaluating its added value in existing risk stratification models.

Many multichannel MCG devices require highly controlled environments, such as magnetically shielded rooms, which restrict their widespread adoption. However, advancements in portable MCG technology, such as those using magnetoresistive sensors, offer the potential for bedside assessments without the need for extensive shielding [17, 41]. Practical improvements in device footprint, usability, operator training, and reduced dependence on shielded environments will be critical factors in facilitating the seamless integration of MCG into clinical practice.

While MCG shows great promise as a tool in post-MI risk assessment, further research is needed to establish standardised diagnostic criteria and optimise clinical implementation. With ongoing technological advancements and larger-scale validation studies, MCG has the potential to become an essential tool for early identification of high-risk post-MI patients, enabling timely intervention and reducing rates of sudden cardiac death.

### 4.1. Limitations

Although these studies demonstrated the potential of MCG as a clinical diagnostic and prognostic tool for post-MI VT patients, several limitations remain. First, there is heterogeneity in study designs, methodologies, diagnostic thresholds, and analytical parameters across the existing literature. Establishing uniform guidelines for MCG interpretation and standardised protocols are crucial for advancing the field. Second, most clinical studies are single centre with small cohorts, which may introduce selection bias and lead to overestimation of diagnostic accuracy. Therefore, larger, multi-center studies involving diverse populations and settings are needed to validate the reproducibility and reliability of MCG. A major barrier to advancing MCG as a clinical tool lies in the limited accessibility of high-quality, large-scale datasets. Currently, the majority of published studies are conducted by a small number of research teams with access to highly specialised, often expensive, MCG equipment, typically located in controlled environments such as magnetically shielded rooms. This restricts both the geographic and methodological diversity of research efforts, and significantly slows progress toward robust, generalisable diagnostic criteria and software tools.

### 4.2. Future trends

To accelerate the development of MCG-based diagnostic methods, particularly those involving complex signal analysis, machine learning, and artificial intelligence, there is a pressing need for large, open-access datasets. By making raw and processed MCG data publicly available, researchers across disciplines, including those without direct access to MCG hardware, can contribute to algorithm development, feature extraction, signal enhancement, and clinical validation. Such democratisation of data access would encourage broader collaboration, spark innovation, and help bridge the gap between engineering-driven methods and clinical translation.

Moreover, open datasets would allow for more rigorous benchmarking of new algorithms, fostering reproducibility and transparency across studies. This is especially important for machine learning approaches, which require diverse and representative datasets to develop models that are both accurate and generalisable to new populations. In addition, access to large-scale MCG data could support the training of early-career researchers and attract interest from data science communities who may not otherwise engage with this field.

Ultimately, to fully realise the clinical potential of MCG, collaborative, data-driven approaches must be supported through investments in shared infrastructure, open science, and cross-disciplinary partnerships. Making high-quality MCG datasets publicly available is a foundational step toward this goal.

Once large-scale MCG datasets become publicly available, the field can begin to shift away from reliance on hand-crafted, expert-defined features toward more modern and data-driven methodologies. Traditional approaches often focus on extracting specific indices, such as QRS duration, RMS amplitude, or fragmentation scores, based on prior physiological knowledge. While informative, these handcrafted features may overlook subtle patterns in the MCG signals that are difficult to define a priori.

With abundant time-series data, researchers can harness powerful deep learning models, such as temporal convolutional networks (TCNs), recurrent neural networks (RNNs), or transformers, to automatically learn relevant features directly from raw or minimally processed signals. These approaches can capture complex temporal dynamics, multi-channel interactions, and non-linear patterns that traditional methods may miss. This paradigm shift enables faster discovery of novel MCG biomarkers, improves classification accuracy, and reduces human bias in feature selection. Ultimately, data-driven methodologies offer the potential for more robust, generalisable, and clinically useful diagnostic tools.

Looking ahead, the vision for MCG extends far beyond specialised hospital settings. With emerging technologies such as optically pumped magnetometers (OPMs) and tunnelling magnetoresistance (TMR) sensors, compact and low-cost systems could enable magnetic field monitoring outside shielded rooms. This opens the possibility of deploying MCG in community clinics or even directly in patients’ homes. Remote, contactless monitoring of cardiac activity in the comfort of the home could revolutionise post-MI care, enabling early detection of arrhythmias and timely intervention. By bringing MCG to everyday environments, we can shift towards more proactive, personalised, and accessible cardiovascular health management.

## 5 Conclusion

MCG presents a non-invasive, contactless, and highly sensitive modality with the potential to improve diagnosis in patients at risk of ventricular arrhythmias following MI. However, further clinical studies are needed to establish a standardised diagnostic criterion and optimize its integration into clinical practice. Additionally, validating and standardizing MCG analytical techniques and parameters for post-MI arrhythmia evaluation is crucial. Finally, based on the existing literature, MCG may have broader clinical applications in diagnosing various types of arrhythmias. Therefore, future clinical studies should explore additional endpoints, such as localizing arrhythmia sources.

## Author Contributions

NGA, MM, and KN each made significant contributions to the conception and design of the study, data acquisition, or data analysis and interpretation. All authors were actively involved in drafting the manuscript or revising it critically for important intellectual content. They have all reviewed and approved the final version for publication. Each author has contributed sufficiently to the work to take public responsibility for relevant sections of the content and has agreed to be accountable for all aspects of the work, ensuring that any issues related to the accuracy or integrity of any part of the study are thoroughly investigated and resolved.

## Ethics Statement

The authors have nothing to report.

## Data Availability Statement

The authors have nothing to report.

